# Temporal Trends in COVID-19 associated AKI from March to December 2020 in New York City

**DOI:** 10.1101/2021.01.18.21249414

**Authors:** Sergio Dellepiane, Akhil Vaid, Suraj K Jaladanki, Ishan Paranjpe, Steven Coca, Zahi A Fayad, Alexander W Charney, Erwin P Bottinger, John Cijiang He, Benjamin S Glicksberg, Lili Chan, Girish Nadkarni

**Affiliations:** Mount Sinai Clinical Intelligence Center, Icahn School of Medicine at Mount Sinai, New York, NY, USA; The Hasso Plattner Institute of Digital Health at Mount Sinai, Icahn School of Medicine at Mount Sinai, New York, NY, USA; Charles Bronfman Institute for Personalized Medicine, Icahn School of Medicine at Mount Sinai, New York, NY, USA; Division of Nephrology, Icahn School of Medicine at Mount Sinai, New York, NY, USA; BioMedical Engineering and Imaging Institute, Icahn School of Medicine at Mount Sinai, Icahn School of Medicine at Mount Sinai, New York, NY, USA; Department of Genetics and Genomic Sciences, Icahn School of Medicine at Mount Sinai, New York, NY, USA; The Pamela Sklar Division of Psychiatric Genomics, Icahn School of Medicine at Mount Sinai, New York, NY, USA; The Department of Psychiatry, Icahn School of Medicine at Mount Sinai, New York, NY, USA; Digital Health Center, Hasso Plattner Institute, University of Potsdam, Professor-Dr.-Helmert-Strasse 2-3, Potsdam, Germany

**Keywords:** COVID-19, Acute Kidney Injury, Renal Replacement Therapy, Hemodialysis

## Abstract

Acute Kidney Injury (AKI) is among the most common complications of Coronavirus Disease 2019 (COVID-19). Throughout 2020 pandemic, the clinical approach to COVID-19 has progressively improved, but it is unknown how these changes have affected AKI incidence and severity. In this retrospective analysis, we report the trend over time of COVID-19 associated AKI and need of renal replacement therapy in a large health system in New York City, the first COVID-19 epicenter in United States.

---

Acute kidney injury (AKI) and need for acute renal replacement therapy (RRT) are frequent complications of Coronavirus Disease-19 (COVID-19) due to SARS-CoV2 infection. During the first surge of the pandemic in the United States, AKI incidence was up to 40% in hospitalized patients and >50% in patients admitted to the intensive care unit (ICU) (1,2). Among patients with COVID-19 related AKI, 17-19% (32% in ICU) required RRT and in-hospital mortality was as high as 50%, with an adjusted odds ratio of almost ten-fold compared to non-AKI patients (1,2). In the following months, multiple therapies have emerged to treat SARS-CoV2 infection. Additionally, our understanding of disease course has advanced, and physician familiarity in managing hospitalized patients with COVID-19 has improved. In particular, high dose corticosteroids have been proven effective in randomized clinical trials (3) and other treatments are supported by growing evidence. (4) These improvements are reflected by the decrease in overall mortality rate for COVID-19 positive AKI patients (5). In this evolving landscape, we sought to investigate temporal trends of AKI over 2020 in the overall COVID-19 hospitalized population and in clinically relevant subgroups from a large healthcare system in New York City; the initial site of the COVID-19 outbreak in the United States.

We analyzed data from the electronic health records (EHRs) from five major hospitals belonging to the Mount Sinai Health System; we included patients with age >18 years, PCR-confirmed SARS-CoV-2 infection and admitted for >48 hours. We excluded patients with known end stage kidney disease (ESKD) on dialysis and kidney transplant recipients. Our primary outcome of interest was AKI defined according to creatinine-based KDIGO criteria. The baseline creatinine was defined as the most recent value obtained within 7-365 days prior to admission or, when unavailable, imputed on the basis of a Modification of Diet in Renal Disease (MDRD) eGFR of 75 ml/min as done previously (1). We identified acute dialysis by procedure codes and nursing flow sheets.

In the first three months of the New York COVID-19 outbreak (from March to May 2020), AKI incidence was consistently >40%; subsequently, it halved to 21.8% in June and remained relatively stable until December (17.4%). Similarly, the proportion of patients needing dialysis also decreased from 20.1% in March vs. 6.2% in December 2020. (**Figure 1**)

**Figure 1.**
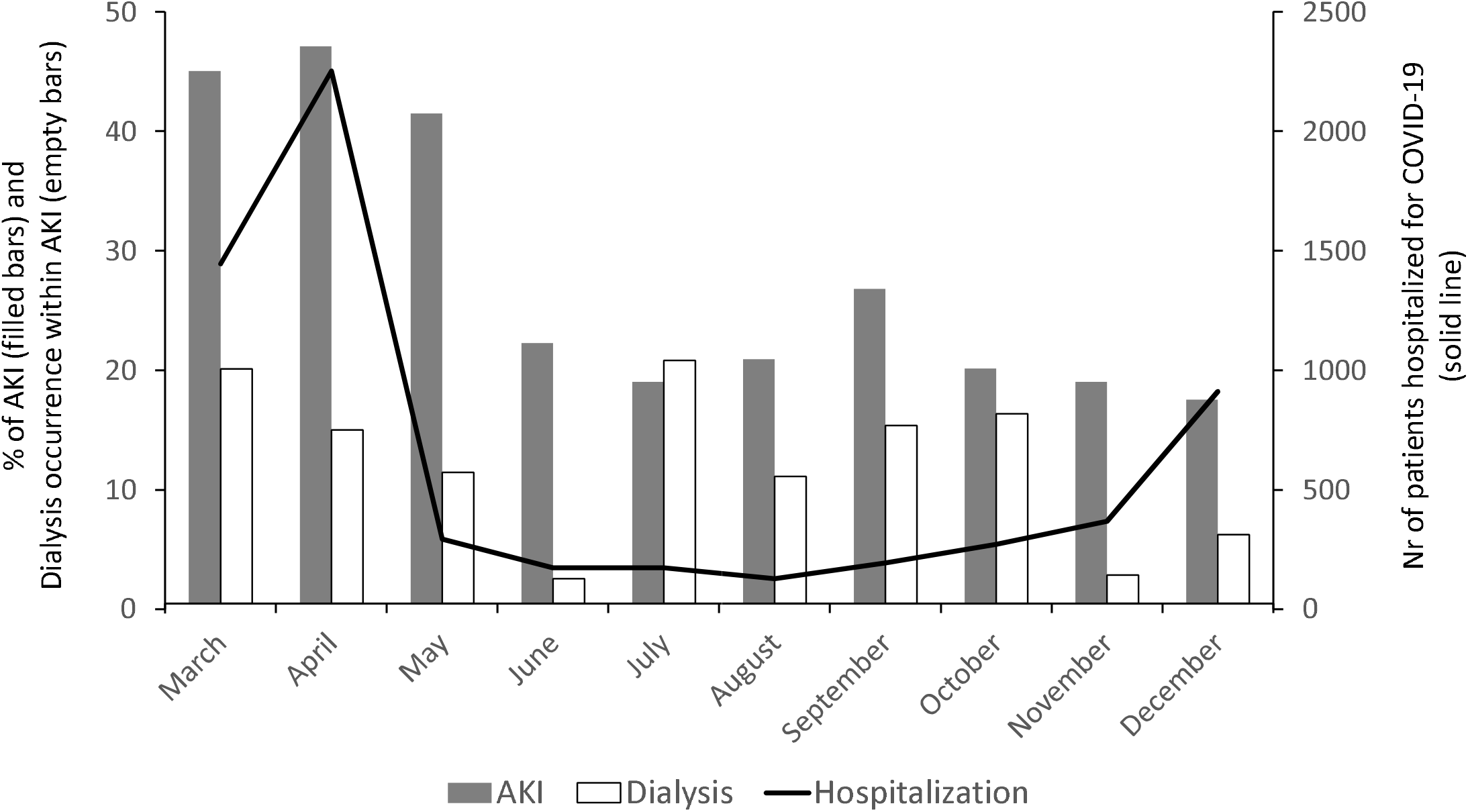
Trends of acute kidney injury incidence through the SARS-CoV2 pandemic in 2020: Bar graphs depict the percentage of patients suffering from acute kidney injury (AKI – filled bars) within those hospitalized for Coronavirus Disease’19 (COVID-19) throughout 2020, and the percentage of patients requiring renal replacement therapy (RRT – empty bars) within the AKI group. The solid line depicts the total number of COVID-19 hospitalizations from March to December 2020.

The reduction in AKI incidence over time was consistent in all subgroup analysis including gender, age and race. AKI incidence in males decreased from a maximum of 50.2% in April to a nadir of 14.1% in November; in female patients the peak was 45.7% in March and the lowest value was 17.2% in December. Of note, the increased incidence of AKI reported in males at the beginning of the pandemic (1) did not persist in the subsequent months. In CKD patients, AKI was observed in >80% of cases in the first two months vs. <40% in the last two. As expected, AKI was more frequent in the oldest subjects, affecting more than 60% of patients older than 75 years in April vs. 23.7% in December. Consistently, all the other age groups (<55yo, 56-65, and 66-75) had a similar halving of AKI incidence. Finally, after stratifying by self-reported race, we observed that African American patients had the highest incidence of COVID-19 AKI throughout the pandemic; the peak value was 51.2% in March which decreased to 23.7% in December 2020. In white patients, the incidence was 45.4 and 13.6% respectively.

In summary, using data from approximately one year of the COVID-19 pandemic in a large, urban health care system, we show that AKI incidence in COVID-19 patients has substantially decreased overall and in key clinically relevant subgroups (age, race, gender and CKD). However, AKI still continues to be a common complication in hospitalized patients with COVID-19 with one in five being affected.

Multiple reasons might explain the observed difference between the first three months and the rest of the pandemic. With the first surge, the health system was partly unprepared to face an event of these proportions and SARS-CoV2 testing availability was not sufficient to estimate the real size of the outbreak (6). Thus, patients hospitalized in the first three months were likely sicker and were managed in a relative lack of human and hospital resources. Second, it is possible that the therapeutic approach to COVID-19 patients has improved in the last months, preventing incidences of AKI. Third, it is also likely that physicians have grown significant experience and are better equipped for the management of COVID-19. Nevertheless, the incidence of AKI is still high in COVID-19; CKD patients have the highest risk of AKI, and racial disparities remain. Continued investigation is needed to further explore AKI in COVID-19 and to assess whether any of the newer therapeutic and management strategies have a causal role in preventing AKI in COVID-19.

## Data Availability

Data are available upon request to the authors

